# The Contribution of Health Behaviours to Occupational Class Inequalities in Cardiovascular Disease: Mediation and Residual Effects

**DOI:** 10.64898/2026.04.06.26349958

**Authors:** O Pietiläinen, L Vähäsarja, A Etholén, E Teppo, J Boch, P Speyer, P Jousilahti, J Harkko, T Lallukka

## Abstract

**Background:** Cardiovascular diseases (CVD) are more common in lower occupational classes, but the mediating role of health behaviours remains unclear. This study aimed to quantify the extent to which health behaviours mediate the association between occupational class and CVD, evaluate their relative contributions to CVD risk, and assess occupational class differences in the effects of health behaviours.

**Methods:** Municipal employees from Helsinki, aged 40–60 at baseline, were followed from 2000–2002 (response rate 67%) to 2022. CVD events were identified from national registers, including hospitalizations, long-term sickness absence, disability pensions, and mortality. Counterfactual mediation analysis using additive survival regression was used to assess the contribution of health behaviours — excessive alcohol consumption, smoking, unhealthy diet, and insufficient physical activity — to the association of occupational class and CVD. Occupational class differences in the effects of health behaviours were assessed with Cox regression.

**Results:** During follow-up, 50% of participants in the low occupational class and 46% in the high occupational class had a CVD event. All unhealthy behaviours except heavy alcohol use were more common in the low occupational class. Health behaviours explained approximately 40% of the excess risk of CVD when moving from high occupational class to low occupational class. Insufficient physical activity (HR 1.44, 95% CI 1.35–1.54) was the strongest predictor of CVD. Unhealthy diet was more strongly associated with CVD in the high occupational class.

**Conclusion:** Health behaviours explained a part of occupational class inequalities in CVD, but most of the inequality remained unexplained, highlighting broader social determinants.

**What is already known:** Occupational class inequalities in cardiovascular diseases are well established, with higher incidence in lower occupational classes. Previous studies suggest that only around one fourth of the association is mediated through health behaviours, but careful quantification through causal mediation methods is needed. Furthermore, few previous studies have examined whether the harmful effects of health behaviours differ by occupational class, which could be an overlooked explanation for inequalities.

**What this study adds:** Using counterfactual mediation analysis, this study suggests that health behaviours mediate around 40% of the negative effect of moving from high to low occupational class on cardiovascular disease. The relative effect of unhealthy diet was stronger in the high occupational class.

## Introduction

Cardiovascular diseases (CVD) have remained a leading cause of death worldwide [1]. Key modifiable risk factors for cardiovascular disease include health behaviours such as unhealthy diet, smoking, excessive alcohol consumption, and insufficient physical activity [2], [3]. Engaging in favourable health behaviours has been associated with a reduced CVD risk of roughly one-half to two-thirds [4], [5], [6].

Individuals in lower occupational classes have consistently been found to experience higher rates of CVD [7]. A major question is how much of this inequality is explained by health behaviours. A previous systematic review suggests that health behaviours account for only around 26% of the association between occupational class and CVD [8]. Understanding how much of the occupational class–CVD gradient is explained by health behaviours is essential for designing interventions and policies. If large proportions of health inequalities remain unexplained by health behaviours, broader social determinants must be targeted.

Health behaviours and other CVD risk factors and protective factors are unequally distributed among socioeconomic groups. Individuals from lower socioeconomic positions tend to have poorer diets, consume more alcohol, smoke more, and exercise less [9]. These differences may explain the occupational class difference in CVD risk.

In addition, if the effects of health behaviours on CVD vary across occupational classes, that may in part explain occupational class differences in CVD. A healthy lifestyle has been found to be more strongly linked to better cardiovascular health in lower socioeconomic positions [10], as well as less cardiovascular mortality in lower socioeconomic positions [11]. The detrimental effects of smoking on morbidity and self-rated health have been found to be weaker in higher socioeconomic positions [12], and similar patterns of alcohol consumption have been found to be more detrimental in lower socioeconomic positions [13] [14].

The primary aim of this study was to investigate the extent to which the association between occupational class and CVD is mediated by health behaviours, and how much remains unexplained by them. Additional aims were to evaluate the relative contributions of occupational class and individual health behaviours to CVD risk and assess whether the effects of health behaviours vary between occupational classes in a longitudinal cohort of current and retired public sector employees in Finland, followed for over two decades.

## Methods

### Study population

The study used a cohort of municipal employees of the City of Helsinki, Finland, from the Helsinki Health Study project. The participants of Phase 1 were followed in repeated surveys. The cohort consisted of 40, 45, 50, 55, and 60-year-old employees of the City of Helsinki in 2000–2002. The participants received baseline questionnaires in 2000–2002 (N=8960, response rate 67%) and were followed up in 2007 (N=7332, response rate 83%), 2012 (N=6814, response rate 79%), and 2017 (N=6832, response rate 82%). The respondents who declined register linkage, those with a cardiovascular event before the baseline questionnaire, or those with missing information on socioeconomic position or health behaviours were excluded from the study, yielding an analytic sample of 5993 participants (73% women).

### Outcome

CVD events were identified from administrative national registers, with ICD-10 codes [15] I00 to I99 identified as cardiovascular disease in all registers. The register sources were: 1) cardiovascular hospitalization (The Finnish Institute for Health and Welfare’s Care Register for Health Care, including data on patients who have been discharged from inpatient care, undergone day surgeries, or received specialized outpatient care), 2) long-term sickness absence spells (12 or more days) (the Social Insurance Institution of Finland), 3) deaths (the Statistics Finland’s Causes of Death Register), and 4) disability pensions (The Disability Pension Register managed by the Finnish Centre for Pensions).

### Occupational class

Occupational class was based on the occupational title from the employer’s personnel register data at the time of answering the Phase 1 questionnaire. Occupational class was categorized into two hierarchical classes, with 1) higher occupational class consisting of managers and professionals and semi-professionals, and 2) lower occupational class consisting of routine non-manual employees and manual workers [16].

### Health behaviours

Leisure-time physical activity was quantified using metabolic equivalent (MET) [17], with individuals with less than 14 MET hours per week classified as having insufficient physical activity [18].

Excessive alcohol consumption was evaluated based on participants’ self-reported weekly alcohol intake, with more than seven weekly units (12 grams of pure ethanol) for women and 14 units for men classified to indicate excessive alcohol consumption [19].

Unhealthy diet was measured by consumption of fruits and vegetables, which was assessed by a food frequency questionnaire over the previous four weeks. Eating fruits and vegetables less than daily was identified as an unhealthy diet.

Smoking status was categorized based on self-reports, distinguishing between current smokers (those who reported using tobacco products daily or occasionally) and non-smokers (those who had never used or had quit using tobacco products).

### Metabolic risk factors

Information on metabolic risk factors was obtained from the questionnaire. Diabetes, hypertension and hypercholesterolemia statuses were based on questions whether the participant has each of these conditions diagnosed by a physician, with answers “Yes” or “No”. Obesity was measured by body mass index (BMI), with a BMI over 30 categorized as obesity.

### Statistical analysis

The participants were followed beginning from the first questionnaire in 2000-2002 until the first CVD event, or until censoring at death for non-CVD causes or the end of year 2022. Cox regression models were used to estimate Hazard ratios and 95% confidence intervals, with the participant’s age used as the time scale.

Health behaviours were treated as time-dependent covariates, changing at each questionnaire where the participant participated and keeping the previous value otherwise. The follow-up was censored after age 75, as the results were inconsistent at older ages, while there was no sufficient data to analyse the effect separately in this age group.

First, Cox regression models were used to estimate the associations of occupational class, each health behavior, sum of the health behaviours, having any of the unhealthy behaviours, and sex, with CVD hazard. The relative contributions of occupational class and health behaviours were examined by calculating an approximation of a Bayes factor comparing the Bayesian information criterion (BIC) of a Cox regression model with each individual health behaviour as an explanatory factor (HBM) to a model with occupational class as an explanatory factor (OCM), and taking a base 10 logarithm of the Bayes factor, with the formula for the Bayes factor being exp((BIC_ocm_ – BIC_hbm_)/2). Bigger values indicate that the model with the health behaviour explains the outcome better than the model with occupational class as an explanatory factor, with values 1/2 to 1 indicating substantial support, 1 to 2 indicating strong support, and 2 or more indicating decisive support for the health behaviour model over the occupational class model according to the interpretation guidelines of Kass & Raftery [20].

To assess the extent to which health behaviours mediate the association of occupational class with CVD, we used an Aalen additive model [21] [22] and the counterfactual procedure introduced by Lange et al. [23]. The total effect of occupational class on CVD is decomposed into the indirect effect, or the effect mediated by the health behaviours, and the direct or residual effect, which is the effect not explained by health behaviours. In practice this is accomplished by estimating the direct effect in the counterfactual situation where each participant would have kept their health behaviours the same and changed only their occupational class, and the indirect effect in the counterfactual situation where each participant would have kept their occupational class the same and changed their health behaviours to the health behaviours of the other occupational class. See Appendix 1 for details. To generate the counterfactual scenarios, we modelled the probability of each of the four mediating health behaviours with binomial logistic regression, adjusting for occupational class, sex, and age.

The occupational class differences in the effects of the health behaviours on subsequent CVD risk were analyzed using Cox proportional hazards regression models. Separate models were fitted for each health behaviour, with CVD being the outcome and the health behaviours, occupational class and their interaction as predictors. The models were stratified by occupational class to allow separate baseline hazards for the occupational classes.

Occupational class-specific HRs and overall marginal HRs averaged over occupational classes were calculated from the models using the emmeans package [24], along with 95% confidence intervals. The p-values of the interaction terms for occupational class and each health behaviour from the models were used to assess the statistical significance of the occupational class difference in the effect of the health behaviour. All analyses were conducted in R 4.3.1 [25].

## Results

This study included 5993 participants, of which 2870 (48%) had a cardiovascular event during the 22-year follow-up (median observed time-at-risk 5.0 years, reflecting the high incidence of CVD events, including sickness absences) (Table 1). The participants were three-fourths women and were divided relatively equally between the high and low occupational classes.

**Table 1:** The number of participants and CVD events by occupational class.

| **Occupational class** | **N** | **No event** | **Event** | **% with event** |
| --- | --- | --- | --- | --- |
| High | 3122 | 1700 | 1422 | 46 |
| Low | 2871 | 1423 | 1448 | 50 |
| Total | 5993 | 3123 | 2870 | 48 |

Cardiovascular events were more common in the low occupational class, with 50% having had an event compared to 46% in the high occupational class. The difference in CVD incidence between occupational classes was statistically significant, with p-value < 0.001 from Chi square test.

Of all participants, 16% of participants smoked, 16% used alcohol excessively, a fifth had an unhealthy diet, and a fifth had insufficient physical activity averaged across the questionnaires (Table 2). Smoking, unhealthy diet, and insufficient leisure-time physical activity were more common among the low occupational class, while excessive alcohol consumption was more common among the high occupational class. Among the participants who had some unhealthy health behaviours, around 60% had only one unhealthy behaviour, around 30% two unhealthy behaviours, around 10% three unhealthy behaviours, and one to two percent had four or five unhealthy behaviours (Supplementary table 1).

**Table 2:** The distributions of unhealthy behaviours by occupational class.

| **Smoking** | **N** | **No** | **Yes** | **% smoking** |
| --- | --- | --- | --- | --- |
| High occupational class | 13254 | 11869 | 1385 | 10 |
| Low occupational class | 11359 | 8894 | 2465 | 22 |
| Total | 24613 | 20763 | 3850 | 16 |

| **Excessive alcohol use** | **N** | **No** | **Yes** | **% excessive alcohol use** |
| --- | --- | --- | --- | --- |
| High occupational class | 13254 | 10865 | 2389 | 18 |
| Low occupational class | 11359 | 9873 | 1486 | 13 |
| Total | 24613 | 20738 | 3875 | 16 |

| **Unhealthy diet** | **N** | **No** | **Yes** | **% unhealthy diet** |
| --- | --- | --- | --- | --- |
| High occupational class | 13254 | 11124 | 2130 | 16 |
| Low occupational class | 11359 | 8673 | 2686 | 24 |
| Total | 24613 | 19797 | 4816 | 20 |

| **Insufficient exercise** | **N** | **No** | **Yes** | **% insufficient exercise** |
| --- | --- | --- | --- | --- |
| High occupational class | 13254 | 10499 | 2755 | 21 |
| Low occupational class | 11359 | 8437 | 2922 | 26 |
| Total | 24613 | 18936 | 5677 | 23 |

The associations of occupational class and health behaviours with CVD and their relative contributions to CVD risk were analyzed with Cox regression models (Table 3). Low occupational class was associated with a higher risk of CVD (HR 1.12, 95% CI 1.06–1.19).

**Table 3:** Contributions of occupational class and unhealthy behaviors to CVD risk, HR and 95% CI, Bayesian Information Criterion (BIC), base 10 logarithm of Bayes factor between the occupational class model and health behaviour models, from Cox regression models.

| Predictor | HR (95% CI) | BIC | Base 10 logarithm of the Bayes factor (occupational class, predictor) |
| --- | --- | --- | --- |
| Occupational class | 1.12 (1.06, 1.19) | 74698 |  |
| Any health behaviour | 1.31 (1.24, 1.38) | 74636 | 13.5 |
| Sum of health behaviours | 1.16 (1.13, 1.20) | 74621 | 16.7 |
| Smoking | 1.16 (1.07, 1.25) | 74698 | 0.0 |
| Excessive alcohol use | 1.02 (0.95, 1.11) | 74712 | -3.0 |
| Unhealthy diet | 1.22 (1.14, 1.31) | 74680 | 3.9 |
| Insufficient physical activity | 1.44 (1.35, 1.54) | 74591 | 23.2 |

Having at least one unhealthy health behaviour had a hazard ratio of 1.31 (95% CI 1.24–1.38), while each additional health behaviour was associated with an increase in risk of HR 1.16 (95% CI 1.13–1.20). Among individual behaviours, insufficient physical activity showed the strongest association with CVD (HR 1.44, 95% CI 1.35–1.54), followed by unhealthy diet (HR 1.22, 95% CI 1.14–1.31) and smoking (HR 1.16, 95% CI 1.07–1.25). Excessive alcohol use was not associated with increased CVD risk (HR 1.02, 95% CI 0.95–1.11). Comparing the relative contributions of occupational class and health behaviours to CVD risk, physical activity and unhealthy diet were stronger predictors of CVD risk than occupational class (decisive support according to the Bayes factor interpretation guidelines), whereas smoking and occupational class predicted CVD risk equally. Occupational class predicted CVD better than excessive alcohol use. Having any one unhealthy behaviour was associated with CVD more strongly than occupational class was associated with CVD, as did any additional unhealthy behaviour (decisive support).

To assess the extent to which the association of occupational class with CVD is mediated through health behaviours, we fitted a counterfactual Aalen additive survival model (Table 4). Transitioning from high to low occupational class would increase the number of CVD events by approximately 27 events per 1000 person years, of which 11 events (40%) mediated through health behaviours, and 16 cases (60%) through other pathways. For moving from low to high occupational class, the total effect is negligible, as the direct and indirect effects operated in opposite directions and largely cancelled each other out.

**Table 4:**
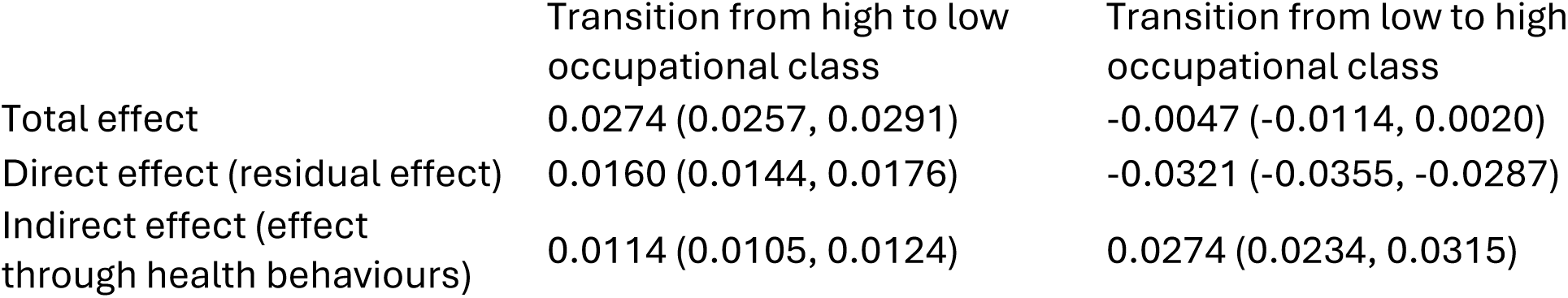
The contrafactual effect of moving between occupational classes in the Aalen model, change in number of cases over one year.

|  | Transition from high to low occupational class | Transition from low to high occupational class |
| --- | --- | --- |
| Total effect | 0.0274 (0.0257, 0.0291) | -0.0047 (-0.0114, 0.0020) |
| Direct effect (residual effect) | 0.0160 (0.0144, 0.0176) | -0.0321 (-0.0355, -0.0287) |
| Indirect effect (effect through health behaviours) | 0.0114 (0.0105, 0.0124) | 0.0274 (0.0234, 0.0315) |

Next, we assessed whether the associations of each health behaviour with CVD risk differed between occupational classes (Table 5). Overall, the associations between health behaviours and subsequent CVD risk were largely similar across the occupational classes, with a statistically significant difference observed only for unhealthy diet. The adverse effect of unhealthy diet was slightly stronger in the higher occupational class (HR 1.32, 95 % CI 1.19 to 1.46 versus 1.13, 95 % CI 1.03 to 1.24 in the lower occupational class; p-value for interaction < 0.01). The association of excessive alcohol use with CVD was slightly stronger in the lower occupational class (HR 1.12, 95 % CI 0.99 to 1.26) than in the higher occupational class (HR 0.98, 95 % CI 0.89 to 1.09), although this difference did not reach statistical significance. The effects of smoking and insufficient exercise were similar across occupational classes.

**Table 5:** The associations of health behaviours with subsequent CVD event by occupational class, HR and 95 % CI.

| HR and 95 % CI for the association of each health behaviour with subsequent CVD |  |  |  |  |
| --- | --- | --- | --- | --- |
| Occupational class | Smoking | Excessive alcohol use | Unhealthy diet | Insufficient physical activity |
| Higher | 1.15 (1.02–1.30) | 0.98 (0.89–1.09) | 1.32 (1.19–1.46) * | 1.44 (1.32–1.58) |
| Lower | 1.13 (1.02–1.24) | 1.12 (0.99–1.26) | 1.13 (1.03–1.24) * | 1.42 (1.30–1.55) |
| All together | 1.14 (1.05–1.23) | 1.05 (0.97–1.13) | 1.22 (1.14–1.31) | 1.43 (1.35–1.53) |

\* p for interaction of occupational class and diet = 0.029

To elucidate the possible biological pathways between occupational class and CVD, we calculated the prevalence of metabolic CVD risk factors in occupational classes (supplementary table 3). Obesity was more common in the lower occupational classes (23% in the low occupational classes vs 16% in the high occupational classes), and diabetes and hypertension were slightly more common in the lower occupational classes (9% vs 7% and 38% vs 34% in the higher and lower occupational classes respectively). There were no occupational class differences in hypercholesterolemia. Metabolic risk factors were not adjusted for, as they form a part of the causal pathway from occupational class to CVD.

## Discussion

CVD was more common in the low occupational class than in the high occupational class. We found health behaviours to explain around 40% of the increased risk of CVD when moving from high occupational class to low occupational class, while the remaining 60% of the association was attributable to other pathways. However, when moving from low to high occupational class, the health behaviour-mediated effects and effects through other pathways acted in opposite directions, leading to a negligible total effect.

Overall, occupational class, smoking, unhealthy diet and insufficient physical activity were associated with increased risk of CVD, while no statistically significant association was found for excessive alcohol consumption. The strongest association was found for insufficient physical activity. It should be borne in mind that alcohol consumption was generally low in this predominantly female cohort, and heavy drinkers are likely underrepresented in the originally employed cohort, which may have attenuated the association between alcohol consumption and CVD.

The detrimental effects of unhealthy behaviors on CVD were largely similar across occupational classes, but unhealthy diet was more detrimental in the higher occupational class. In addition, except for excessive alcohol use, unhealthy behaviors were more prevalent among individuals in the lower occupational class, indicating a higher disease risk in this group.

Previous studies have shown that those in lower socioeconomic positions tend to have poorer health behaviours [9], and this was also confirmed in our study. Similarly to the findings of our study, the differences in health behaviours between occupational classes have been found to explain around 26% of occupational class inequalities in CVD [8]. The differences in health behaviours may themselves be explained by occupational class differences in physical or financial resources or in perceptions of the benefits of healthy behaviours or the harms of negative health behaviours [8].

In our study most health behaviours explained CVD risk more than occupational class alone. However, this is expected, since health behaviours mediate the association between occupational class and CVD. Since a significant part of that association remained unexplained by health behaviours, it remains essential to address broader social determinants of CVD.

Different effects of the health behaviours in occupational classes could provide an additional explanation to the observed differences in CVD, in addition to the more commonly assessed explanation of occupational class differences in the distributions of the explanatory factors. Previous studies have not assessed whether there are socioeconomic differences in the effects of health behaviors on CVD. In our study the associations of health behaviours with subsequent CVD were largely similar between high and low occupational classes, although we did find a more negative effect of unhealthy diet on CVD among the high occupational class. This finding is partly in accordance with previous studies. Zhang et al. [10], found that in lower socioeconomic groups, a healthy lifestyle was more strongly associated with subsequent cardiovascular incidence in a UK population, while no socioeconomic differences were found in a US population. The associations of healthy lifestyle and all-cause mortality, cardiovascular disease incidence, and cardiovascular mortality have also been observed to be stronger in lower socioeconomic positions [11]. Similarly, the detrimental effects of smoking on morbidity and self-rated health have been observed to be stronger in lower occupational classes [12], a finding which was not corroborated by our study. Similarly, Birch et al. [26] observed the harmful effects of smoking on self-rated health to be stronger among those with low income or who were unemployed, whereas it was observed to be less harmful among those with low education.

Although the association of alcohol consumption and CVD was not statistically significant in this cohort, we did observe a more negative effect of excessive alcohol consumption on CVD among the low occupational class, although the occupational class difference was not statistically significant. Nevertheless, the direction of effect aligns with previous studies, which found that harmful effects of alcohol consumption were stronger in lower socioeconomic positions [13] [14]. Although the socioeconomic differences in the harmful effects of alcohol use may reflect differences in alcohol usage patterns, the previous studies have found the socioeconomic difference to persist even when differences in usage patterns are accounted for. Given the overall low level of alcohol consumption and the likely underrepresentation of heavy drinkers in the originally employed cohort, these results should be interpreted with caution.

Our supplementary analyses showed that there are occupational class differences in some of the metabolic risk factors, particularly obesity and diabetes, with the risk factors being more common in the low occupational class. Although no detailed causal conclusions can be drawn from this, this suggests that the occupational class-CVD-link is likely to follow the common metabolic pathways of development of CVD.

### Strengths and limitations of the study

Certain factors lend credibility to our results. First, we have a comprehensive set of outcomes from national registers with a long follow-up. All the outcome measures are based on a physician’s diagnosis and, therefore, are free from possible self-reporting bias. Being from administrative registers, they are also not subject to bias caused by non-participation. However, our registers don’t necessarily capture all cardiovascular events, for example, sickness absences shorter than 12 days.

A weakness of our study is that there were a selection of the sample and a loss of follow-up, which appeared to be partly selective. Attrition was higher among those with worse health behaviours and those in lower socioeconomic positions (supplementary table 2). All study participants were followed for the outcomes on the registers until the end of 2022, but if they dropped out of the follow-up questionnaires, their health behaviour status was not updated in the follow-up questionnaires and reflected earlier exposures. This selection may cause an underestimation of the detrimental effects of poor health behaviours, as those with poor health behaviours are more likely to drop out, and their health behaviour status reflects their health behaviours before dropping out. While this may cause some bias to the results, the selective attrition doesn’t appear to be so strongly biased as to have significantly altered the main findings.

Our study used a longitudinal design to measure health behaviours before cardiovascular illness, which was observed in the registers. This gives credibility to a causal interpretation of the association of health behaviours and the CVD outcome but nevertheless cannot completely rule out the possible effect of selection. Suppose early symptoms of CVD are present when answering the questionnaires. In that case, the participant may have altered their health behaviours already before the occurrence of diagnosed illness, for example, by reducing physical activity before a diagnosis of CVD.

Occupational class, used in this study, reflects many significant aspects of the person’s working conditions and of the person’s socioeconomic position more generally. However, it may not capture all aspects of socioeconomic position as well as other measures, such as wealth, which may be a better indicator of access to resources, or education, which may be significant especially for health behaviors. Therefore, the results should be interpreted as concerning mainly occupational class, and the associations regarding other measures of socioeconomic position warrant further study.

It should be born in mind that a vast majority of the participants were women, who may have less CVD or have undiagnosed CVD, and exhibit a different profile of health behaviours than men. The study participants were also employed at the beginning of the follow-up and therefore excluding persons too ill to work. Therefore, our results are likely to reflect the situation of women better than men and employed populations better than the full population.

### Conclusion

Our results suggest that health behaviours mediate around 40% of the negative effect of moving from high occupational class to a low occupational class on CVD. Smoking, unhealthy diet, and insufficient physical activity were associated with subsequent CVD risk, with insufficient physical activity having the strongest association. Unhealthy diet was more detrimental in the high occupational class. Public health programmes aiming to reduce cardiovascular disease should therefore target physical inactivity, diet and smoking, but also address broader social factors that drive occupational class inequalities in cardiovascular disease.

## Acknowledgements

None.

## Funding

The Novartis Foundation supported this study. The Research Council of Finland (Grant #330527) supported OP, ET, and TL.

## Conflict of interest

Johannes Boch and Peter Speyer were employed by the funding organization, Novartis Foundation, and their involvement in the study should be considered in light of this relationship.

## Data availability statement

The Helsinki Health Study survey data cannot be shared due to strict data protection laws and regulations. More details are available at project website (https://www.helsinki.fi/en/researchgroups/helsinki-health-study) and in our data protection statement (helsinki.fi/en/researchgroups/helsinki-health-study/data-protection-statement). More information about the data can be requested at. Permission to the secondary use of health and social data is granted by the Finnish Social and Health Data Permit Authority, Findata.

## Authors’ Contributions

**Olli Pietiläinen:** Conceptualization, Methodology, Formal Analysis, Investigation, Data Curation, Writing - Original Draft, Writing - Review & Editing. **Luka Vähäsarja:** Conceptualization, Methodology, Software, Formal Analysis, Investigation, Data Curation, Writing - Original Draft, Writing - Review & Editing. **Antti Etholén**: Writing - Review & Editing. **Eero Teppo**: Writing - Review & Editing. **Johannes Boch:** Conceptualization, Funding Acquisition, Writing - Review & Editing. **Peter Speyer:** Conceptualization, Funding Acquisition, Writing - Review & Editing. **Pekka Jousilahti:** Writing - Review & Editing. **Jaakko Harkko:** Data Curation, Writing - Review & Editing. **Tea Lallukka:** Conceptualization, Funding Acquisition, Project Administration, Resources, Supervision, Writing - Review & Editing.

## Supplements

**Supplementary table 1:** Cumulation of unhealthy behaviours.

|  | % with 0 bad<br>health<br>behaviours | % with 1 bad<br>health<br>behaviours | % with 2 bad health<br>behaviours | % with 3 bad health<br>behaviours | % with 4 bad health<br>behaviours |
| --- | --- | --- | --- | --- | --- |
| Smoking: no | 0.59 | 0.32 | 0.08 | 0.01 | 0.00 |
| Smoking: yes | 0.00 | 0.36 | 0.40 | 0.20 | 0.04 |
| Excessive alcohol use: no | 0.59 | 0.30 | 0.09 | 0.02 | 0.00 |
| Excessive alcohol use: yes | 0.00 | 0.44 | 0.35 | 0.17 | 0.04 |
| Unhealthy diet: no | 0.61 | 0.30 | 0.07 | 0.01 | 0.00 |
| Unhealthy diet: yes | 0.00 | 0.42 | 0.37 | 0.17 | 0.03 |
| Insufficient physical activity: no | 0.64 | 0.27 | 0.07 | 0.01 | 0.00 |
| Insufficient physical activity: yes | 0.00 | 0.51 | 0.33 | 0.13 | 0.03 |

**Supplementary table 2:** Attrition analysis, attrition in phases 2-5 according to different explanatory factors (OR, 95% CI).

|  | Phase 1 | Phase 2 | Phase 3 | Phase 4 | Phase 5 |
| --- | --- | --- | --- | --- | --- |
| Missing obs., n (%) | 0 (0.0) | 1020 (15.4) | 1365 (20.6) | 1396 (21.1) | 2016 (30.5) |
| Total | n=6602 |  |  |  |  |
| Men | n=1420 | 1.80 (1.55 - 2.09) | 1.59 (1.39 - 1.82) | 1.48 (1.29 - 1.69) | 1.49 (1.31 - 1.68) |
| Low education (Phase 1) | n=2622 | 1.33 (1.16 - 1.52) | 1.26 (1.12 - 1.43) | 1.48 (1.31 - 1.66) | 1.87 (1.69 - 2.08) |
| Manual labour (Phase 1) | n=1045 | 1.68 (1.42 - 1.98) | 1.58 (1.36 - 1.84) | 1.60 (1.38 - 1.86) | 1.96 (1.72 - 2.25) |
| Isomnia symptoms (Phase 1) | n=1320 | 0.88 (0.74 - 1.04) | 0.78 (0.67 - 0.92) | 0.85 (0.73 - 1.00) | 1.02 (0.89 - 1.16) |
| Above moderate drinking (Phase 1) | n=1101 | 1.21 (1.02 - 1.44) | 1.24 (1.06 - 1.45) | 1.26 (1.08 - 1.46) | 1.22 (1.06 - 1.40) |
| Current smoker (Phase 1) | n=1516 | 1.61 (1.39 - 1.87) | 1.79 (1.57 - 2.05) | 1.77 (1.55 - 2.02) | 1.75 (1.55 - 1.97) |
| Non-daily consumption of F&Vs (Phase 1) | n=1544 | 1.48 (1.28 - 1.72) | 1.56 (1.36 - 1.78) | 1.52 (1.33 - 1.73) | 1.47 (1.30 - 1.65) |
| Physical inactivity (Phase 1) | n=1584 | 1.34 (1.15 - 1.55) | 1.31 (1.15 - 1.50) | 1.31 (1.15 - 1.50) | 1.32 (1.17 - 1.49) |
| Depression / Anxiety (Phase 1) | n=1025 | 1.24 (1.04 - 1.48) | 1.08 (0.91 - 1.27) | 1.37 (1.17 - 1.60) | 1.29 (1.12 - 1.49) |
| RAND general health (Phase 1) | n=1831 | 1.32 (1.14 - 1.52) | 1.31 (1.15 - 1.49) | 1.45 (1.28 - 1.65) | 1.56 (1.39 - 1.75) |
| CV medication (<Phase 1) | n=1631 | 1.10 (0.94 - 1.28) | 1.11 (0.97 - 1.27) | 1.19 (1.04 - 1.36) | 1.36 (1.21 - 1.53) |
| Nervous system medication (<Phase 1) | n=1605 | 1.16 (1.00 - 1.35) | 1.18 (1.03 - 1.35) | 1.20 (1.05 - 1.37) | 1.27 (1.13 - 1.44) |

**Supplementary table 3:** Metabolic CVD risk factors by occupational class, N (%).

|  | Occupational class |  |  |
| --- | --- | --- | --- |
|  | High<br>(N=14442) | Low<br>(N=12719) | Overall<br>(N=27161) |
| Obesity |  |  |  |
| No | 12030 (83.3%) | 9581 (75.3%) | 21611 (79.6%) |
| Yes | 2264 (15.7%) | 2939 (23.1%) | 5203 (19.2%) |
| Missing | 148 (1.0%) | 199 (1.6%) | 347 (1.3%) |
| Diabetes |  |  |  |
| No | 10588 (73.3%) | 8325 (65.5%) | 18913 (69.6%) |
| Yes | 944 (6.5%) | 1208 (9.5%) | 2152 (7.9%) |
| Missing | 2910 (20.1%) | 3186 (25.0%) | 6096 (22.4%) |
| Hypertension |  |  |  |
| No | 8621 (59.7%) | 6904 (54.3%) | 15525 (57.2%) |
| Yes | 4856 (33.6%) | 4784 (37.6%) | 9640 (35.5%) |
| Missing | 965 (6.7%) | 1031 (8.1%) | 1996 (7.3%) |
| Hypercholestrolemia |  |  |  |
| No | 8271 (57.3%) | 6980 (54.9%) | 15251 (56.2%) |
| Yes | 5017 (34.7%) | 4458 (35.1%) | 9475 (34.9%) |
| Missing | 1154 (8.0%) | 1281 (10.1%) | 2435 (9.0%) |

## Appendix 1: Additional details about the counterfactual Aalen additive model

The Aalen additive model is a semiparametric model for survival outcomes assuming consistency, identifiability, and no unmeasured confounders (Aalen, 1980; Martinussen et al. 2006; Lange & Hansen, 2011). Importantly, the model allows for causal interpretation through contrafactual rate differences (for details, see Lange & Hansen, 2011). In the Aalen model, the rate *γ* at time *t* is defined as

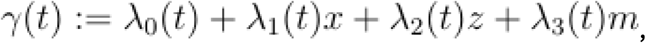

where x is the exposure, m is the mediator, and z is a vector of other baseline covariates. Each λ_j_ (t) is a potentially time-dependent coefficient function, which can be estimated from data. A λ_j_ may also be time-invariant (that is, a constant function), which simplifies estimation (Lange & Hansen, 2011).

Lange et al.’s (2012) method consists of four main steps.

1. Model the exposure X and the mediator M conditional on confounders using the original data.
2. Construct a new dataset with a new variable A* that takes every possible value of x for each participant. The height of the dataset should be n times the original height, when n is the number of levels of x.
3. Compute the weights by applying the models from step 1 to the new data set. In R, this is achieved with the predict function.
4. Fit a new model including only x and A* as covariates but weighted by the weights from step 3. The natural direct and indirect effects may now be derived from this model.

## Notes

### Author Declarations

Ethics Committee of the Faculty of Medicine, University of Helsinki, approved The Helsinki Health Study protocol. The City of Helsinki health and personnel authorities approved The Helsinki Health Study protocol. The Finnish Social and Health Data Permit Authority Findata granted the permission to the secondary use of social and health care data.

